# Efficacy of Jiedu Tongluo Tiaogan Formula on Type 2 Diabetic Lower Extremity Arteriosclerosis Obliterans: Study Protocol for a Metabolomics-Based Randomized Controlled Clinical Trial

**DOI:** 10.1101/2022.06.25.22276893

**Authors:** Tianjiao Liu, Chunli Piao, Yuting Peng, Jinghan Xu, Yue Qu, Qi Li, Xiaohua Zhao, Xuemin Wu, Pei Li, Yawen Fan

**Affiliations:** Institution of Shenzhen Hospital, Guangzhou University of Chinese Medicine (Futian), Shenzhen 518000, Guangdong Province, P.R. China

**Keywords:** Jiedu Tongluo Tiaogan Formula, type 2 diabetic mellitus, lower extremity arteriosclerosis obstruction, randomized controlled trial, metabolomics

## Abstract

**Introduction:** Diabetic lower extremity arteriosclerosis obstruction (DLASO) is a common macrovascular complication in type 2 diabetic mellitus (T2DM), which can cause amputations and a higher risk of cardiovascular events. However, there are few effective treatments for DLASO currently. To evaluate the safety and efficacy of Jiedu Tongluo Tiaogan Formula (JTTF) in type 2 diabetic lower extremity arteriosclerosis obliterans and looking for a mechanism of action, we designed a clinical trial and mechanism exploration based on metabolomics technology.

**Methods and Analysis:** This study is designed as a randomized controlled clinical trial. A total of 80 participants will be recruited and randomized to a TCM group (JTTF + essential treatments) and a control group (essential treatments) in a ratio of 1:1. The treatment duration is 12 weeks. Changes in clinical symptom scores, color ultrasound Doppler hemodynamics of lower extremity arteries, and Ankle-Brachial Ratio (ABI) will be the primary outcomes. Changes in TCM symptoms scores, other indicators related to arteriosclerosis, blood glucose, lipids, and body mass will be the secondary outcomes. The primary and secondary outcomes will be evaluated at baseline and week 12. Safety outcomes and adverse events will also be properly assessed. After treatment completion, blood and urine samples from subjects will be tested for metabolomics.

**Discussion:** This study aims to verify the efficacy and safety of JTTF in type 2 diabetic lower extremity arteriosclerosis obliterans and obtain the key action pathway. It helps to provide scientific evidence for TCM treatment of diabetic vascular complications.

**Ethics and Dissemination:** This trial has been approved by the Ethics Committee (GZYLL(KY)-2021-024). Results of this trial will be published in journals and presented at scientific conferences. We share raw data in the ResMan network platform. All the authors declare that they have no conflicts of interest.

**Trial registration:** Chinese Clinical Trials Register, ChiCTR2100051337. Registered on 20 September 2021.

## INTRODUCTION

Type 2 diabetes mellitus (T2DM) is a chronic metabolic disease often linked to long-term complications. China has the largest population of patients with diabetes worldwide; the prevalence of diabetes among Chinese people aged 18 and over was 11.2%(1). In China, the number of patients with T2DM will be predicted to double in 2030. In economically advanced countries, the increase will be about 50% in 2030(2). The severity of T2DM is that it greatly increases the incidence of cardiovascular disease, cerebrovascular disease, and peripheral macrovascular disease(3). As a large global study shows, microvascular complications accounted for about half the number of diabetic complications (4).

Diabetic lower extremity arteriosclerosis obstruction (DLASO), one of the common macrovascular complications of T2DM, can directly lead to symptoms such as numbness, cold or heat, unbearable pain, and intermittent claudication, etc. Moreover, it can cause lower-extremity ischemic ulcers and amputations, increase the risk of cardiovascular events, and lead to higher mortality (5). It diminishes the health-related quality of life and causes a considerable financial burden to patients. Therefore, exploring the related risk factors and pathogenic mechanisms of type 2 diabetic lower extremity arteriosclerosis obliterans is of great significance, and seeking effective therapeutic drugs.

However, the pathogenesis of DLASO has not been elucidated. The risk factors recognized mainly include age (6), gender (7), smoking, blood sugar(8, 9), blood lipids(10, 11), blood pressure (12), fibrinogen (Fib) (13, 14), serum uric acid (UA) (15), C-reactive protein (CRP) (16), homocysteine (Hcy), cystatin C (CysC) (17). For the mechanism of DLASO, the standard theories include Vascular endothelial injury and smooth muscle cell proliferation theory, lipid infiltration theory, hemodynamic change theory, inflammatory response theory, thrombosis theory, and genetic theory. Presently, there is no specific drug for the treatment of DLASO, and only general hypoglycemic, hypotensive, and lipid-lowering treatments can be given.

Combining Western medicine with traditional Chinese medicine (TCM) has gradually become the primary approach to treating many diseases. TCM possesses advantages based on its pleiotropic, multi-target, prospective, and stable features that enable it to alleviate T2DM and other complex chronic diseases (18). Under the guidance of TCM theory, Chinese patent medicines used alone or in conjunction with western medicine could successfully lower blood glucose levels and relieve symptoms in patients with diabetes and diabetic complications (19, 20). A clinical study confirmed that Chinese patent medicine (Mudan granule) was effective and safe in the treatment of DLASO and also regulated lipids, dissolved Fib, increased blood flow rate, and dilated blood vessels (21).

Jiedu Tongluo Tiaogan Formula (JTTF), a Chinese patent medicine, consisting of Coptis chinensis Franch (Huanglian), Radix Rhei Et Rhizome (Dahuang), Astragalus propinquus Schischkin (Huangqi), Salvia miltiorrhiza Bunge (Danshen), Bupleuri Radix (Chaihu), etc. A series of previous studies have confirmed that JTTF can play a role in treating T2DM by enhancing autophagic flux, protecting islet β cells, restoring cellular homeostasis, and reducing insulin resistance in animal and cell experiments(22-25). Relevant clinical trials have also confirmed that JTTF can improve glucose and lipid metabolism disorders in patients with T2DM and reduce insulin resistance (26-28). However, previous studies of JTTF have involved small sample sizes and without clinically registered information. Moreover, there is still a lack of clinical research on diabetes complications, especially the research to verify its mechanism of action through metabolomics technology. For these reasons, high-quality clinical evidence of the safety and efficacy of JTTF in individuals with type 2 diabetic lower extremity arteriosclerosis obliterans is required urgently. To seek an effective therapy for type 2 diabetic lower extremity arteriosclerosis obliterans and to provide new methods and ideas for the clinical prevention and treatment of the disease, we conduct a clinical trial and mechanism exploration based on metabolomics technology. This study aims at presenting the methodologies and full details of the protocol.

## METHODS AND ANALYSIS

### Study Design

This study is designed as a randomized controlled clinical trial following the Helsinki Declaration and the guidelines for good clinical practice. A total of 80 participants who satisfy the inclusion and exclusion criteria and sign the informed consent form will be randomly divided into two groups, which will both be administered essential treatments, consisting of diabetes education, exercise, diet, as well as rational control of blood glucose, blood pressure, lipids. The TCM group will be received the intervention with essential treatments +JTTF (200 ml twice a day). And the control group will be received only essential treatments. The treatment duration is set at 12 weeks. The data in the experiment will be collected using a case report form (CRF) and statistically analyzed by SPSS. After treatment completion, blood and urine samples from 60 subjects in two groups will be tested for metabolomics (Taking into account the cost of metabolomics testing and the variability of human samples, we select 30 cases in each population group to study the metabolic differences of each group). We hypothesize that JTTF has good efficacy and safety in patients with Type 2 diabetic lower extremity arteriosclerosis obliterans and would like to obtain the key action pathway by analyzing the metabolic changes. The trial schematic flow is illustrated in Figure 1.

**Figure 1.**
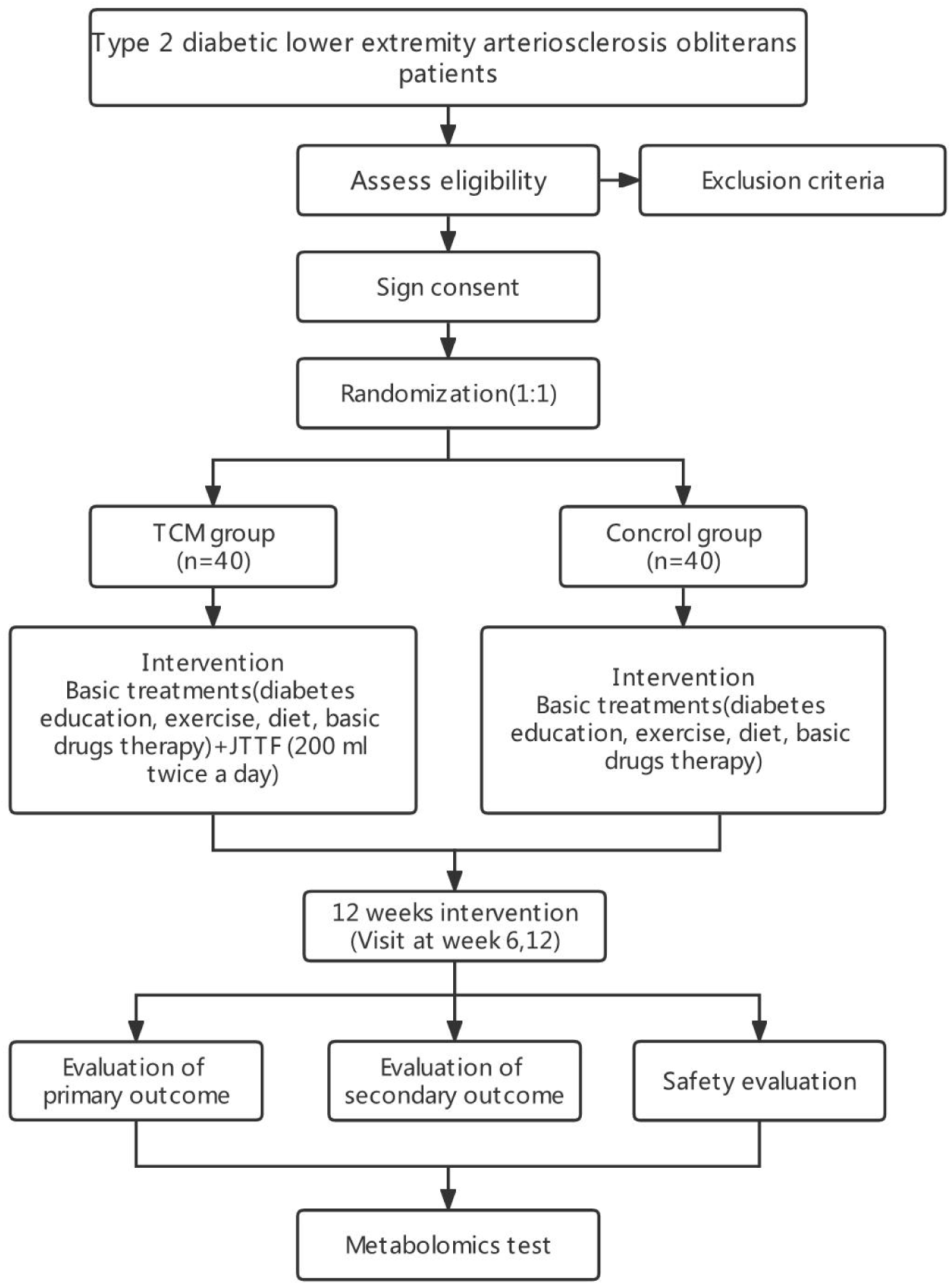
The trial schematic flow

### Research Setting

This study will recruit 80 eligible subjects from Shenzhen Hospital (Futian) of Guangzhou University of Chinese Medicine. For all research diagnoses, blood index, and Color Doppler ultrasound tests, Chinese and Western medicine treatments will be carried out in the Shenzhen Hospital (Futian) of Guangzhou University of Chinese Medicine.

### Inclusion Criteria

1. Meet the diagnostic criteria for type 2 diabetes developed by the American Diabetes Association in 2021(29):
  (1) Fasting plasma glucose≥7.0 mmol/L (126 mg/dl)
    - Fasting is defined as no caloric intake for at least 8 h*
  (2) Two-hour plasma glucose≥11.1 mmol/L (200 mg/dl) during OGTT
    - The OGTT test should be performed as described by WHO using a glucose load containing the equivalent of 75g of anhydrous glucose dissolved in water.*
  (3) HbA1c≥6.5% (48 mmol/mol).
    - The test should be conducted in a laboratory using a method that is National Glycohemoglobin Standardization Program certified and standardized to the Diabetes Control and Complications Trial assay.*
  (4) In patients with typical symptoms of hyperglycemia or hyperglycemic crisis, a random plasma glucose≥11.1 mmol/L (200 mg/dl). * If any of the above four items meet the standard and also meet the standard in repeated testing on the following day, T2DM can be diagnosed.
2. Meet the diagnostic criteria for lower extremity arteriosclerosis obliterans issued by the Vascular Surgery Group of the Chinese Society of Surgery in 2016 (30):
  (1) The age of onset is usually more than 40 years old;
  (2) Have high-risk factors such as smoking, diabetes, hypertension, hyperlipidemia;
  (3) Clinical manifestations of lower extremity arterial occlusive disease, such as intermittent claudication, rest pain, decreased extremity skin temperature, hair loss, etc.;
  (4) The arterial pulse of the distal limbs weakens or disappears;
  (5) ABI≤0.9;
  (6) Color Doppler ultrasound, CTA, MRA, DSA, and other imaging examinations show the stenosis or occlusion of corresponding arteries. * If the first four criteria are met, the clinical diagnosis of ASO can be made. ABI and color ultrasound can determine the degree of ischemia in the lower extremity.
3. According to the Guidelines for Clinical Research of Chinese Medicine (New Drug) (31), a patient with at least two of the primary symptoms and more than two of the secondary symptoms listed below to be diagnosed with Qi deficiency and collaterals obstruction syndrome:
  (1) Primary symptoms and signs: ① numbness; ② pain; ③paresthesia
  (2) Secondary symptoms and signs: ① scaly dry skin; ② dim complexion; ③ fatigue; ④ Shortage of Qi and lackadaisical; ⑤ spontaneous sweating
  (3) Tongue condition: light-dark/petechiae tongue, with thin and white coating.
  (4) Pulse condition: thin and blocked pulse.
4. Aged over 18 years including male and female;
5. Not suffering from severe cardiovascular and cerebrovascular diseases, such as stroke, coronary heart disease, myocardial infarction;
6. Liver and kidney function tests normal;
7. Agree and sign the informed consent form.

### Exclusion Criteria

1. Not meeting the diagnostic criteria of T2DM, lower extremity ASO, and TCM syndrome of Qi deficiency and collaterals obstruction syndrome;
2. Type 1 diabetes mellitus (T1DM) or latent autoimmune diabetes in adults (LADA) patients;
3. Under the age of 18;
4. Patients with comorbidities such as cardiovascular, renal, hepatic, and hematopoietic system and other severe primary diseases, severe coagulation mechanism disorder, mental illness, or other conditions that may affect clinical research as judged by the investigator;
5. Severe intermittent claudication affects the life quality of the patient, and the conservative treatment is not practical, or there is severe ischemia, necrosis, or infection in the limbs, and those who meet the indications for surgery;
6. Patients are pregnant or breastfeeding or planning to become pregnant within 6 months;
7. Severe allergic constitution, known or possible allergy to the test drug or its components;
8. Those who have participated in other clinical trials in the past 3 months.

### Withdrawal Criteria

#### The Withdrawal Decided by the Researcher

The researcher can decide to withdraw a subject from the study under the following circumstances:

1. During the study, the subjects may develop severe complications or pathological changes (such as allergy, abnormal renal and liver function, hospitalization, and other adverse events) that are not appropriate for this study;
2. During the study, the subjects may have poor adherence to medications(for example, the use of drugs does not reach 50% or exceeds 50% of the prescribed dose);
3. Other treatment drugs were added without following the researcher’s guidance during the entire study period.

#### Subjects Withdrawal at Their Own Will

According to the informed consent form, subjects have the right to withdraw from the trial at any point. Subjects who do not formally withdraw from the trial but no longer receive drugs and undergo testing or who are lost to follow-up are also regarded as withdrawn. Understand the reasons for their withdrawal as much as possible and record them(such as perceived poor curative effect, intolerable to some adverse reactions, and economic factors).

Case record forms (CRF) should be kept for cases withdrawn from the study, regardless of the reason.

#### Randomization

Statisticians not involved in the experiment will be asked to create a table of random numbers on a computer. Patients successfully enrolled in this study will be assigned to either the treatment or control groups in a ratio of 1:1. The random serial number will be sealed in an opaque envelope and provided only to clinicians so that participants can be assigned treatment plans accordingly. Since this trial is an open-label study, the participants and physicians will be aware of the allocation protocol, and data analysts and statisticians will not know the distribution results.

#### Sample Size Calculation

Based on the previous study (28), JTTF reduced HbA1c levels in T2DM patients with liver and stomach heat accumulation syndrome after 12 weeks of treatment; the treatment group was 6.94±0.67, and the control group was 7.50±0.92, and the mean difference between these two groups was 0.56. Considering the type I error of 5% (α = 0.05) and a power of 90% (1-β= 0.9, β= 0.1), the experimental group and the control group were allocated 1:1. The sample size was calculated by the Power Analysis and Sample Size (PASS) software (version 15.0). A total of 64 patients will be needed. According to the dropout rate of 20%, we set the final sample size as 80, with 40 patients in each group, taking into account the number of dropout samples.

#### Interventional Methods

All included subjects will accept essential treatment, including diabetes education, exercise, diet, as well as rational control of blood glucose, blood pressure, and lipids, according to the 2020 diagnosis and treatment guidelines for T2DM in China (1) and the 2021 American Diabetes Association guidelines (29). Subjects will be randomly allocated to either the TCM group (JTTF, including Coptis chinensis Franch (Huanglian), Radix Rhei Et Rhizome (Dahuang), Astragalus propinquus Schischkin (Huangqi), Salvia miltiorrhiza Bunge (Danshen), Bupleuri Radix (Chaihu), etc., 200 ml twice a day) or the control group (only essential treatment). Treatment will continue for 12 weeks.

#### Outcome Assessment

We use the case report form (CRF) to collect demographic Information and efficacy outcomes, including primary and secondary outcomes, safety outcomes, and adverse events.

All efficacy outcomes measurements will be performed at baseline and week 12. Safety outcomes and Adverse Events assessment will be performed at baseline, week 6, and week 12. The time point and study period are shown in Figure 2.

**Figure 2.**
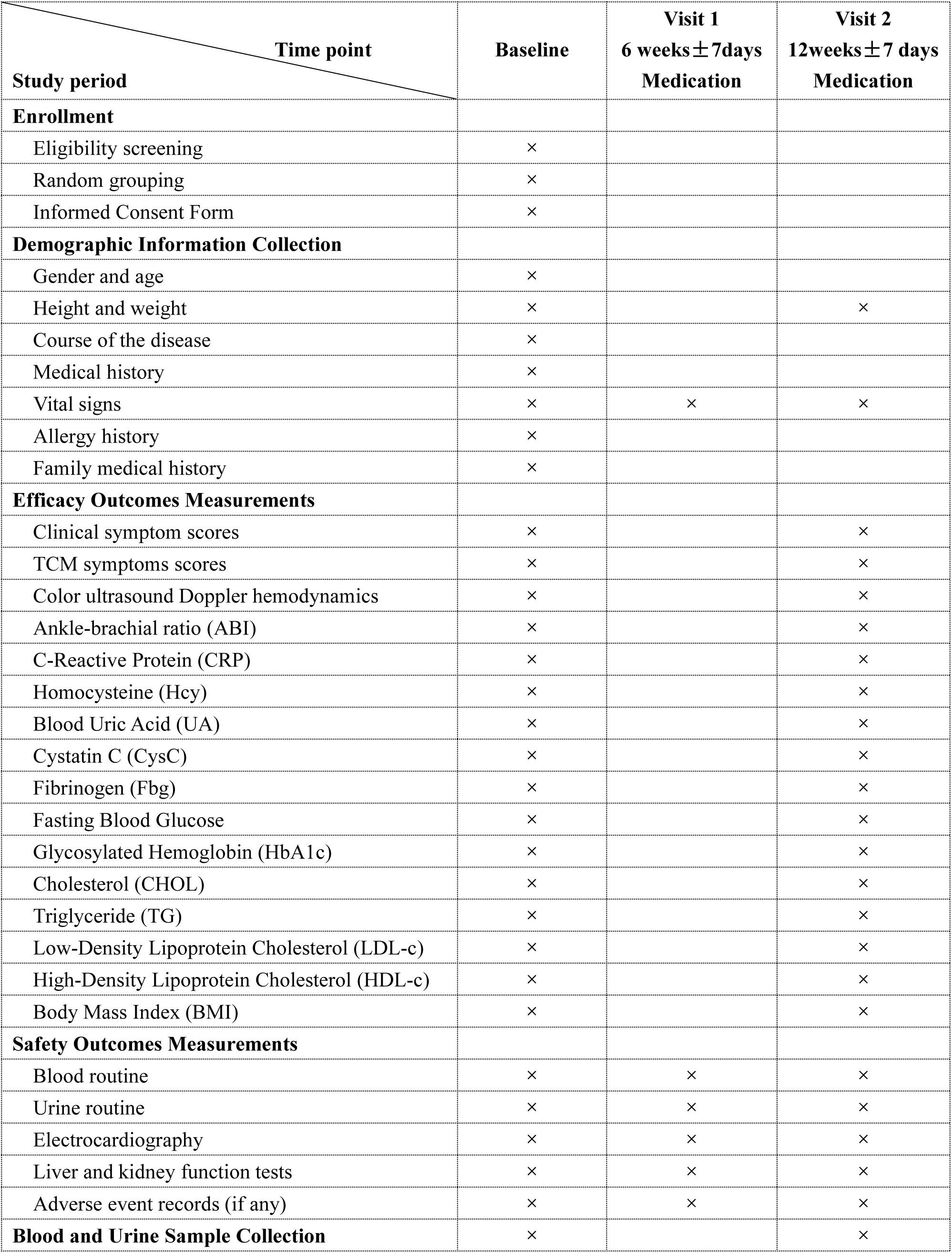
Time Point and Study Period

#### Demographic Information

Demographic information such as gender, age, height, weight, course of the disease, previous medical history, allergy history, family medical history will be collected.

#### Primary Outcomes

The primary outcome of the study is the changes in mainly indicators associated with diabetic atherosclerosis:

1. Changes in clinical symptom scores(Additional file 1, according to The curative effect standard of ASO made by Peripheral Vascular Committee of China Association of Chinese Medicine,2002);
2. Changes in color ultrasound Doppler hemodynamics of lower extremity arteries (Inner diameter of common femoral artery, popliteal artery, anterior tibial artery, and dorsal pedis artery);
3. Changes in Ankle-Brachial Ratio (ABI).

#### Secondary Outcomes

Changes in TCM symptoms scores, other indicators related to arteriosclerosis, and indicators associated with blood glucose, lipids, body mass:

1. Changes in TCM symptoms scores (Additional file 2, according to Guidelines for Clinical Research of Chinese Medicine (New Drug),2002);
2. Changes in other indicators related to arteriosclerosis: C-Reactive Protein (CRP), Homocysteine (Hcy), blood Uric Acid (UA), Cystatin C (CysC), Fibrinogen (Fbg) levels;
3. Changes in indicators associated with blood glucose: Fasting Blood Glucose, Glycosylated Hemoglobin (HbA1c) levels;
4. Changes in indicators associated with lipids: Cholesterol (CHOL), Triglyceride (TG), Low-Density Lipoprotein Cholesterol (LDL-c), High-Density Lipoprotein Cholesterol (HDL-c) levels;
5. Changes in Body Mass Index (BMI) levels.

Clinical symptom scores will be measured with additional file 1, and TCM symptoms scores will be measured with Additional file 2. A professional sonographer will perform a color ultrasound Doppler test and vascular diameter measurement. ABI test and BMI (weight (kg) / Height (m)^2^) measurement will be performed by the endocrinologist. Patients should keep fasting for at least 8 hours before the blood test.

#### Safety Outcomes and Adverse Events

1. Adverse events: adverse events such as diarrhea, nausea, vomiting, severe allergies, hospitalization, or surgery, will be carefully recorded at any time. The intensity of adverse events will be categorized as light, moderate and heavy. Serious adverse events (SAE) must be reported immediately to the hospital’s ethics committee, and the subjects should receive complementary clinical treatment. At the same time, it will be reported to the National Medicinal Product Administration (NMPA) within 24 h.
2. General vital signs (such as body temperature, heart rate, respiration);
3. Blood routine, urine routine, 12-lead Electrocardiography, liver and kidney function tests.

#### Data Analysis

The data collected in the experiment will be statistically analyzed by SPSS(version 24.0). According to the specific values of each index, the Kolmogorov-Smirnov test is used to determine whether it conforms to the normal distribution. Suppose the index conforms to the normal distribution. In that case, the results will be expressed in the form of mean ± standard deviation (X ± S), the independent sample T-test is used to judge the difference between the two groups, and the paired sample T-test is used to judge the difference between each group before and after the intervention. If the index does not conform to the normal distribution, the χ2 test and Fisher ‘s exact test are used to judge the difference between the two groups. Spearman correlation analysis is used to test the correlation of each index. All statistical tests will be two-sided; p < 0.05 will be considered statistically significant. For missing data, we delete them or use methods like multiple imputation for data processing.

### Metabolomics Test

#### Sample Collection

There were 30 healthy control groups (No history of major diseases of the heart, brain, liver, kidney, and other vital organs, no diabetes), 30 type 2 diabetes groups (No lower extremity arteriosclerosis obliterans), 30 TCM groups (JTTF), and 30 control groups, a total of 120 subjects. Blood samples were collected from the subjects, of which the traditional Chinese medicine treatment group and the control group were collected before and after treatment, with a total of 180 samples.

Venous blood (4 ml) and midstream urine (10 ml) samples were collected from each patient after overnight fasting. All samples were collected at approximately the same time each day (between 8 and 10 am every day). The collected blood samples were left standing for 30 minutes at room temperature and then placed in a centrifuge. The specific parameters were a temperature 4°C, a rotation speed of 3000 rpm, and a centrifugation time of 5-10 minutes to separate serum. Pipette an equal volume of supernatant 1ml and place them in marked 1.5ml sterile EP tubes. Immediately store serum and urine samples in a -80°C refrigerator after dispensing.

#### Metabolomics Analysis Process

After the sample was dissolved, add 400μl (methanol solution) to the sample, vortex for 1 minute, mix well, and then centrifuge at 12,000 rpm for 10 minutes (4°C). Take 200 μl of the supernatant and place it in a vacuum centrifugal concentrator to concentrate and drain the water. To be tested, take 20 μl from each sample and mix it to form a QC sample.

After obtaining the raw data, follow the following steps to process the data: First, the raw data of the mass spectrometer needs to be preprocessed, including extracting the peak list information and using the QC sample information based on the actual sample signal. The local polynomial regression fitting signal correction (Quality control–based robust LOESS signal correction, QC-RSC) method was used to process the data. Then, the differentially expressed metabolites under different treatment conditions were found through statistical analysis, and the metabolic pathway analysis was carried out.

#### Patient and Public Involvement statement

In the research, patients and the public are only being participants. We do not have involvement from patients or members of the public in the designor conduct, or reporting, or dissemination plans of the research.

## DISCUSSION

T2DM is one of the most harmful chronic diseases in the 21st century due to its many complications, low cure rate, and management challenges. Nevertheless, T2DM may be treatable with TCM formulations based on highly intuitive principles for achieving clinically curative effects that have shown to be effective in clinical practice.

TCM treats diseases based on the viewpoint that recognizes the interconnectedness of all body systems. In the theory of TCM, DLASO is related to the poor circulation of qi, blood, body fluids, and the obstruction of the meridians. One TCM agent, JTTF, was formulated based on TCM theory. JTTF mainly consists of five herbal medicines, namely Coptis chinensis Franch (Huanglian), Radix Rhei Et Rhizome (Dahuang), Astragalus propinquus Schischkin (Huangqi), Salvia miltiorrhiza Bunge (Danshen), Bupleuri Radix (Chaihu), which are combined in relative proportions by weight of 15:9:15:15:10, respectively. Pharmacological studies confirmed that Huanglian could effectively inhibit gluconeogenesis and promote glycolysis to achieve the effect of lowering blood sugar (32). Dahuang can reduce serum cholesterol and increase high-density lipoprotein, delaying fatty liver (33). Huangqi has a two-way regulation function on blood sugar and has a good effect on the treatment of diabetes and its chronic complications (34). Danshen has anti-platelet aggregation and anti-oxidation effects and has a good effect on the treatment of coronary heart disease(35). Chaihu has good anti-inflammatory, anti-fatty liver, and anti-liver damage effects(36). Results of the previous study suggested that JTTF could inhibit the production of inflammation-inducing factors in a multi-target manner, promote the secretion of adiponectin, reduce the inflammatory response of adipocytes, and alleviate endoplasmic reticulum stress (24). In addition, JTTF can alleviate insulin resistance and reduce apoptosis while further increasing glucose and lipid metabolism in the treatment of T2DM rats (25). However, the efficacy and mechanism of JTTF in treating T2DM complications, especially in DLASO, remain unclear. Thus, designing and conducting a high-quality clinical trial to assess the efficacy and safety and to seek the mechanism of JTTF in treating DLASO is of great value.

This study aims to verify the efficacy and safety of JTTF in patients with type 2 diabetic lower extremity arteriosclerosis obliterans through prospective clinical studies. Meanwhile, we use metabolomics technology to obtain the key action pathway of JTTF in treating type 2 diabetic lower extremity arteriosclerosis obliterans. Metabolomics is an emerging discipline with rapidity, accuracy, high resolution, high sensitivity, and a small number of samples required. Therefore, it is an effective method for discovering disease-related biomarkers(37). In recent years, more and more studies about metabolites determined the possible biomarkers and their metabolic pathways according to their differences, providing an essential theoretical basis for the effective prevention and treatment of T2DM(38). In this study, researchers perform overall profile detection and analysis of endogenous small-molecule metabolite intermediates and end products in biological samples under physiological and pathological conditions. It helps to correlate better the abstract description of TCM with objective biological parameters and provides an overall systematic research method and modern scientific evidence for TCM syndrome differentiation and treatment.

However, this study also has certain limitations. Firstly, this trial was not blinded. Lower extremity arteriosclerosis obliterans increase the risk of diabetic foot and amputation. In order not to delay the treatment of patients, this study did not use a placebo control. Secondly, patients aged younger than 18 years were not included in this research. The occurrence of lower extremity arteriosclerosis obliterans is related to the course of diabetes. Few patients are in this age group. Additionally, considering medication safety for underage patients, we decided to exclude this specific population from the study.

## Supporting information

SPIRIT checklist

## Data Availability

All data produced in the present study are available upon reasonable request to the authors
All data produced in the present work are contained in the manuscript
All data produced are available online at the ResMan network platform (https://www.chictr.org.cn/edit.aspx?pid=134097&htm=4).

## ETHICS AND DISSEMINATION

### Ethics Statement

This trial has been approved by the Ethics Committee (Shenzhen Hospital (Futian) of Guangzhou University of Chinese Medicine). The approval number is GZYLL(KY)-2021-024. Written informed consent for the collection and use of participant data and biological specimens will be obtained from all the participants enrolled. Before signing the informed consent form, the researcher illustrates the procedure, potential risks, and benefits of participating in this research. Researchers will have access to the final trial dataset, and all personal information of participants is kept confidential during and after the trial.

### Dissemination and Consent for Publication

Results of this trial will be published in peer-reviewed journals and presented at scientific conferences. All subjects were asked for permission to publish the study results and assured of anonymity and confidentiality.

### Data Sharing

We share raw data from trials, including technical appendix, statistical code, and study dataset available from the ResMan network platform (https://www.chictr.org.cn/edit.aspx?pid=134097&htm=4).

### Research Monitoring

This study will be conducted under the supervision of the GCP(Good Clinical Practice) Office of Guangzhou University of Traditional Chinese Medicine Shenzhen Hospital (Futian), the GCP office will review the progress of the study, patient enrollment, and funding use on a monthly basis. The process will be independent from investigators and the sponsor.

### Trial Status

The trial was registered with the Chinese Clinical Trials Register (ChiCTR2100051337) on 20 September 2021 (https://www.chictr.org.cn/edit.aspx?pid=134097&htm=4). Patients are being recruited continuously from October 2021 to the present. 50 subjects have completed the 12-week intervention. The follow-up visits are expected to be completed in September 2022. Metabolomics testing will be completed by November 2022. The anticipated start date for the analysis is December 2022 and completed in January 2023.

### Author Contributions

Tianjiao Liu and Chunli Piao contributed equally to this work and are the co-first author. Chunli Piao and Tianjiao Liu conceived and designed the research methods. Chunli Piao obtained funding. Tianjiao Liu and Yuting Peng collected the data and drafted the manuscript. Tianjiao Liu, Jinghan Xu, and Yawen Fan analyzed the data. Qi Li, Xiaohua Zhao, Xuemin Wu, and Pei Li carried out the experimental validation. All authors read and approved the final manuscript.

### Funding

Chunli Piao was supported by the National Natural Science Foundation of China (grant number: 81973813), the Scientific research project of Traditional Chinese Medicine Bureau of Guangdong Province (grant number: 20221329), and Shenzhen Science and Technology Innovation Program (grant number: JCY20190809110015528). The funder was not involved in the study design, collection, analysis, interpretation of data, the writing of this article, or the decision to submit it for publication.

---

**Additional File 1.**
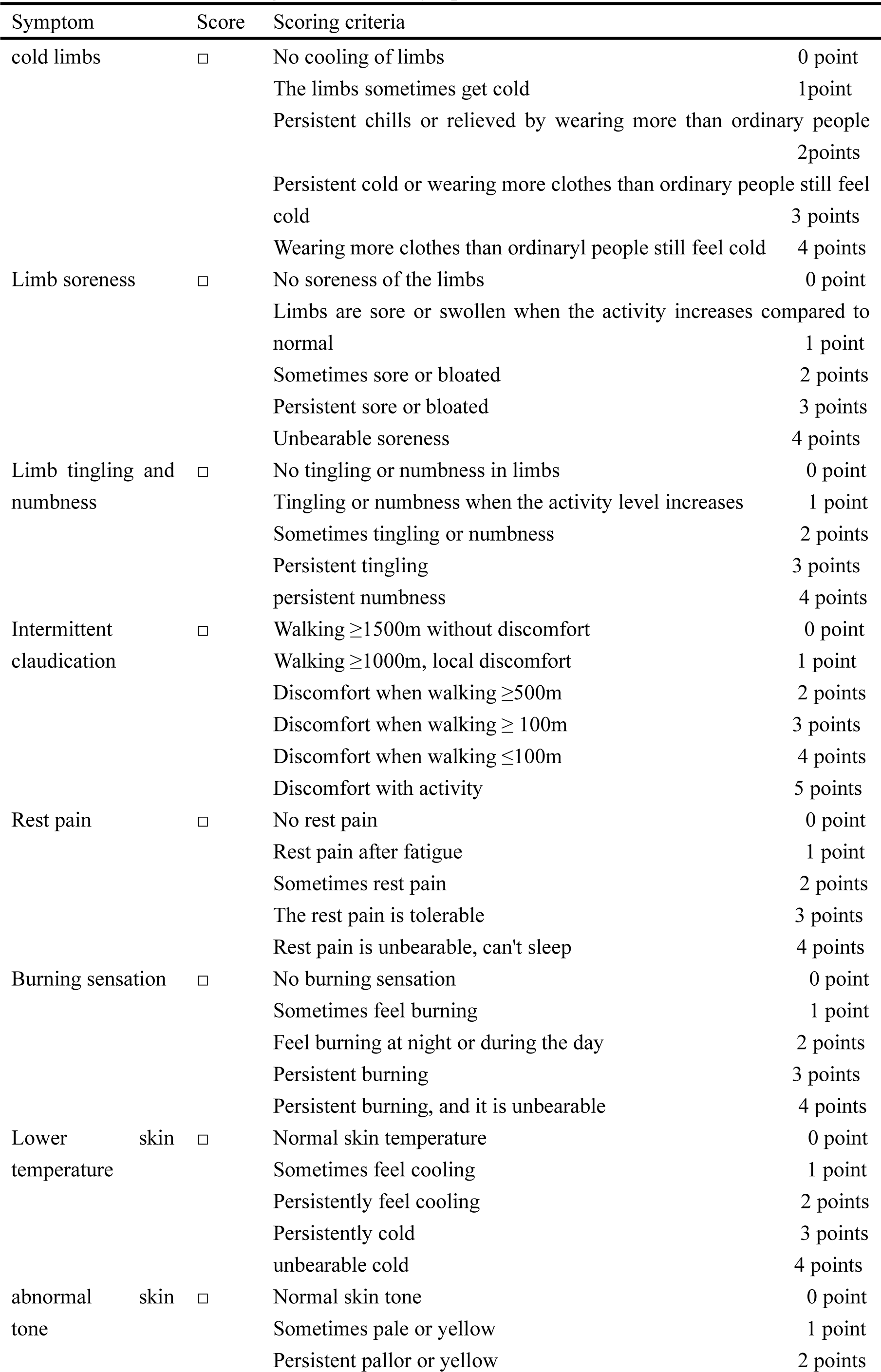
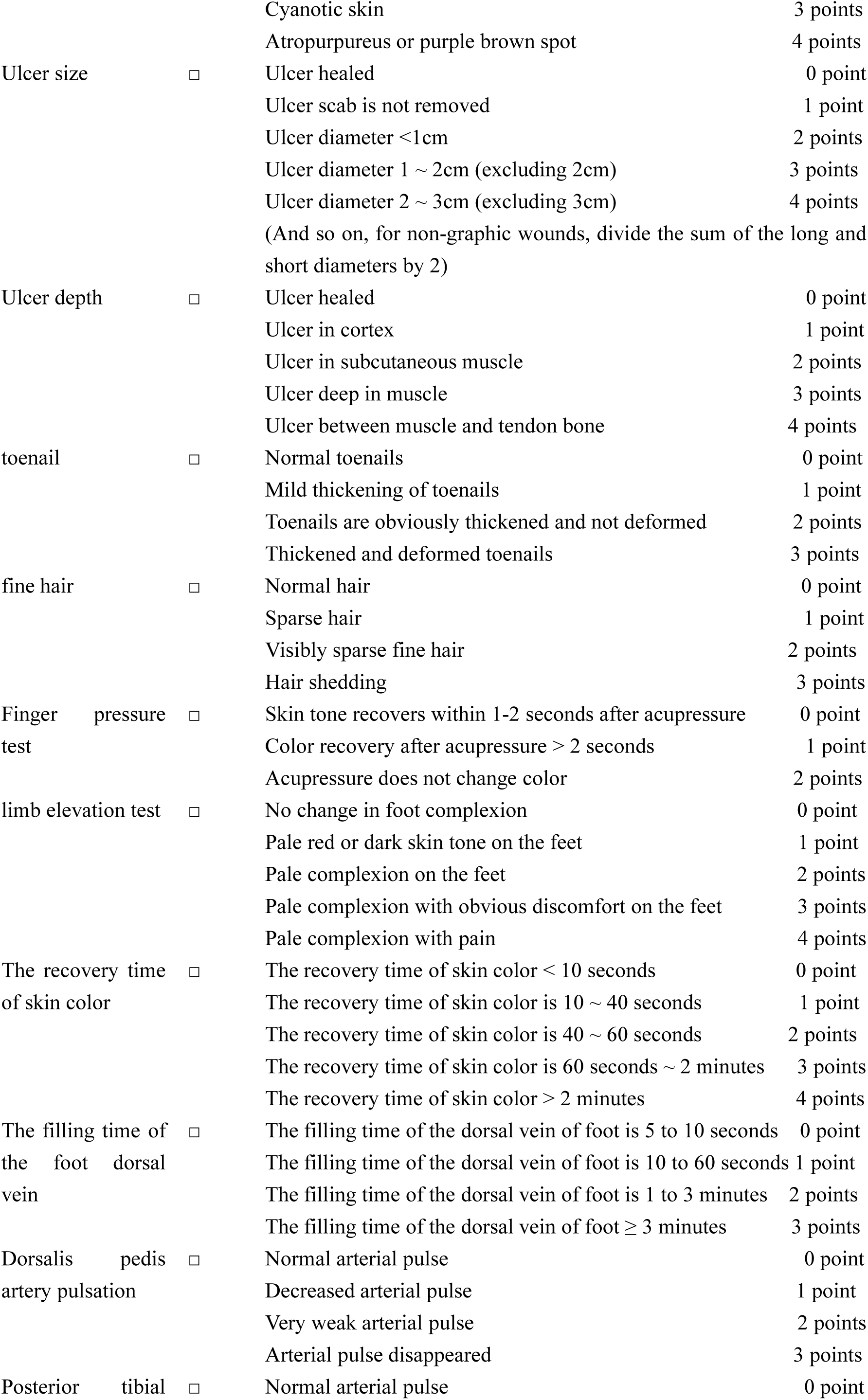
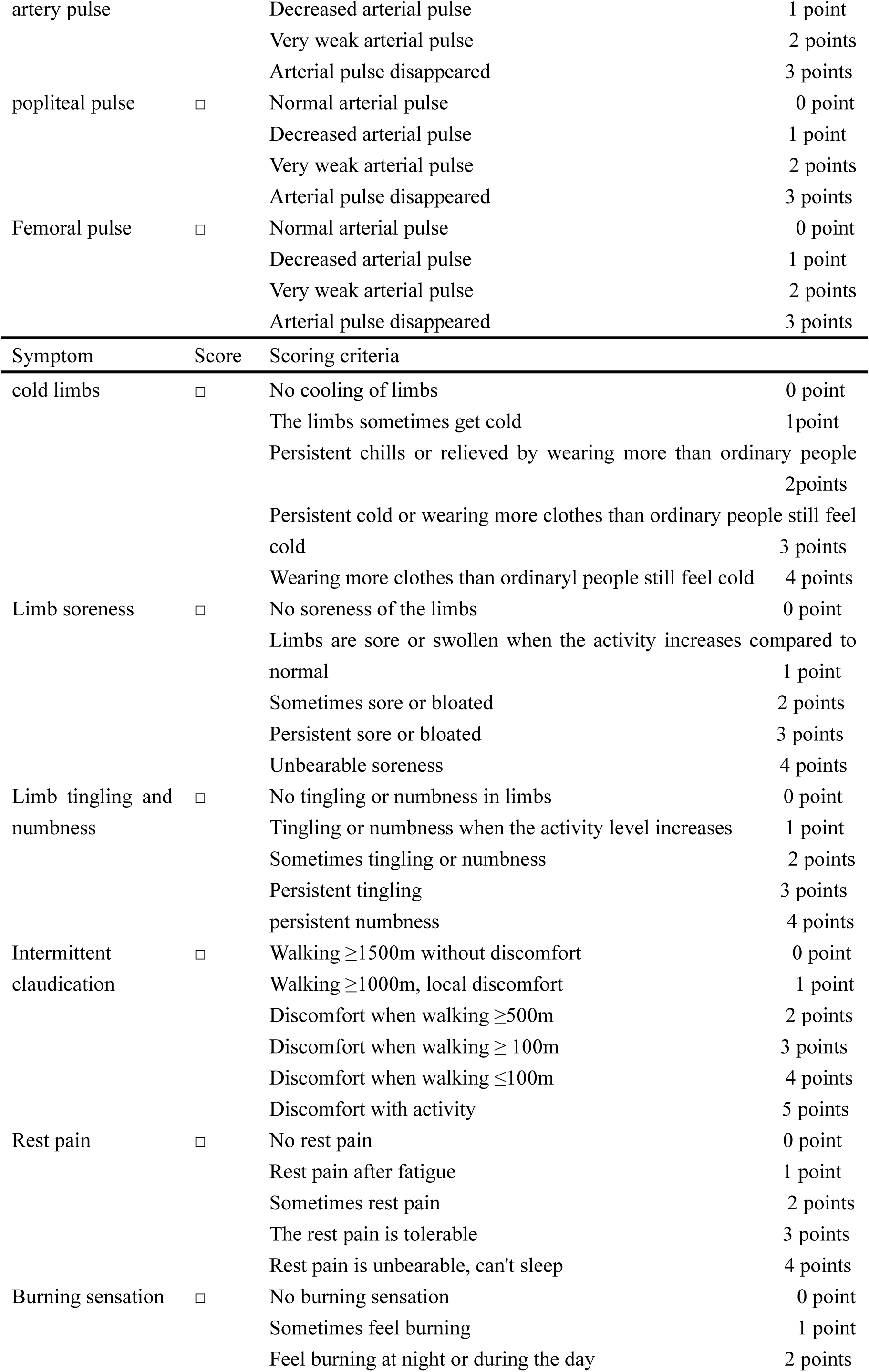
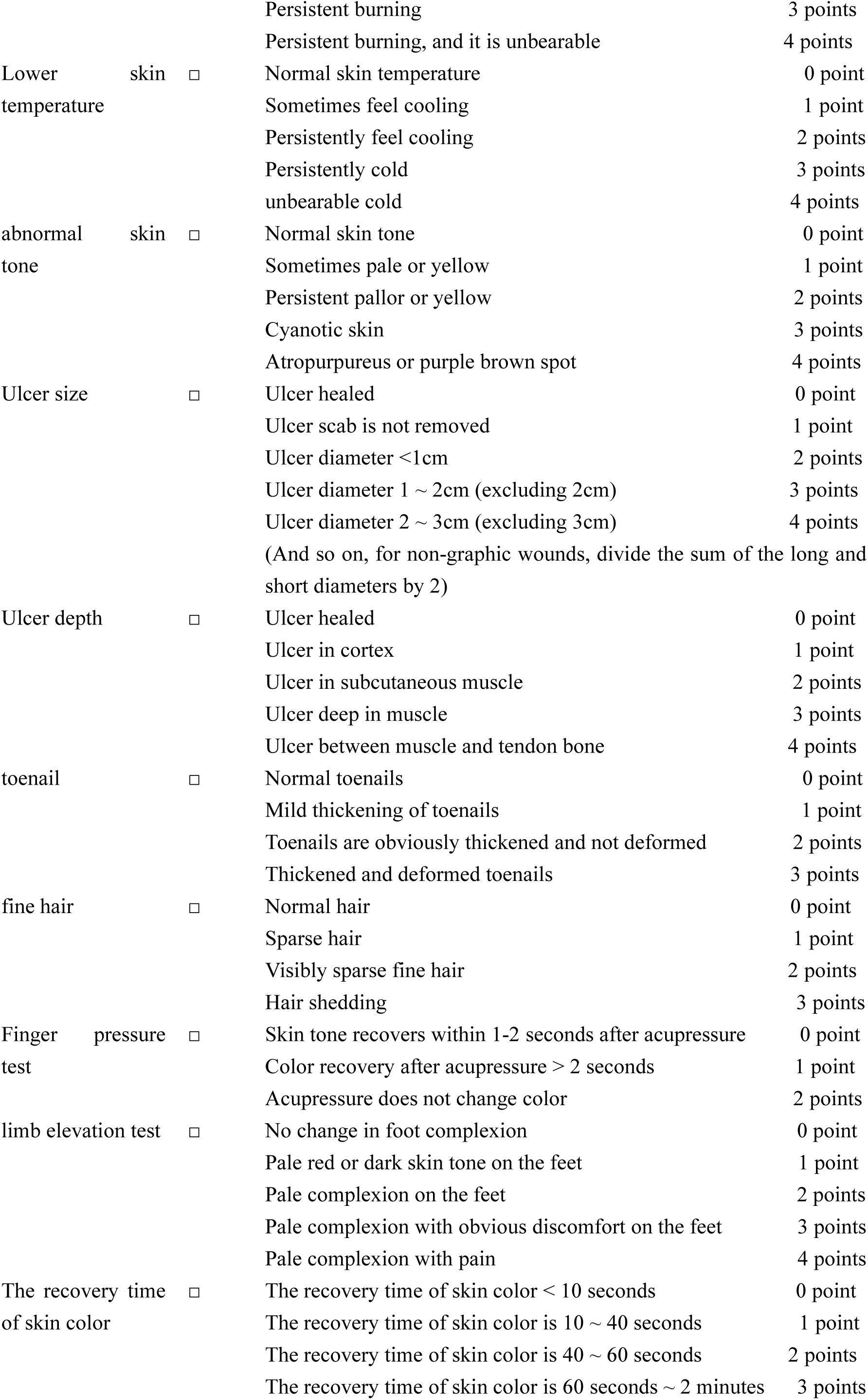
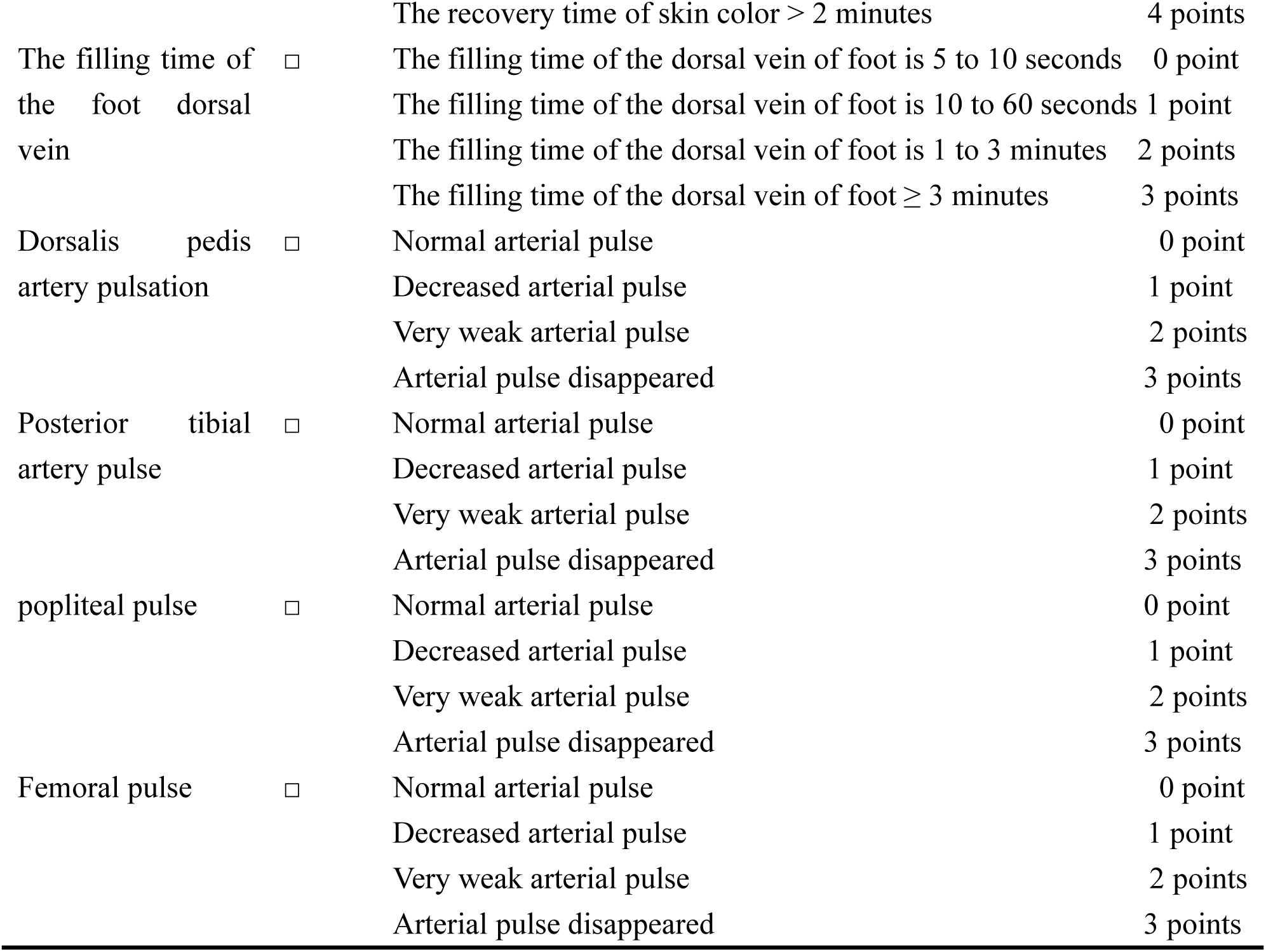
Changes in clinical symptom scores

**Additional File 2.**
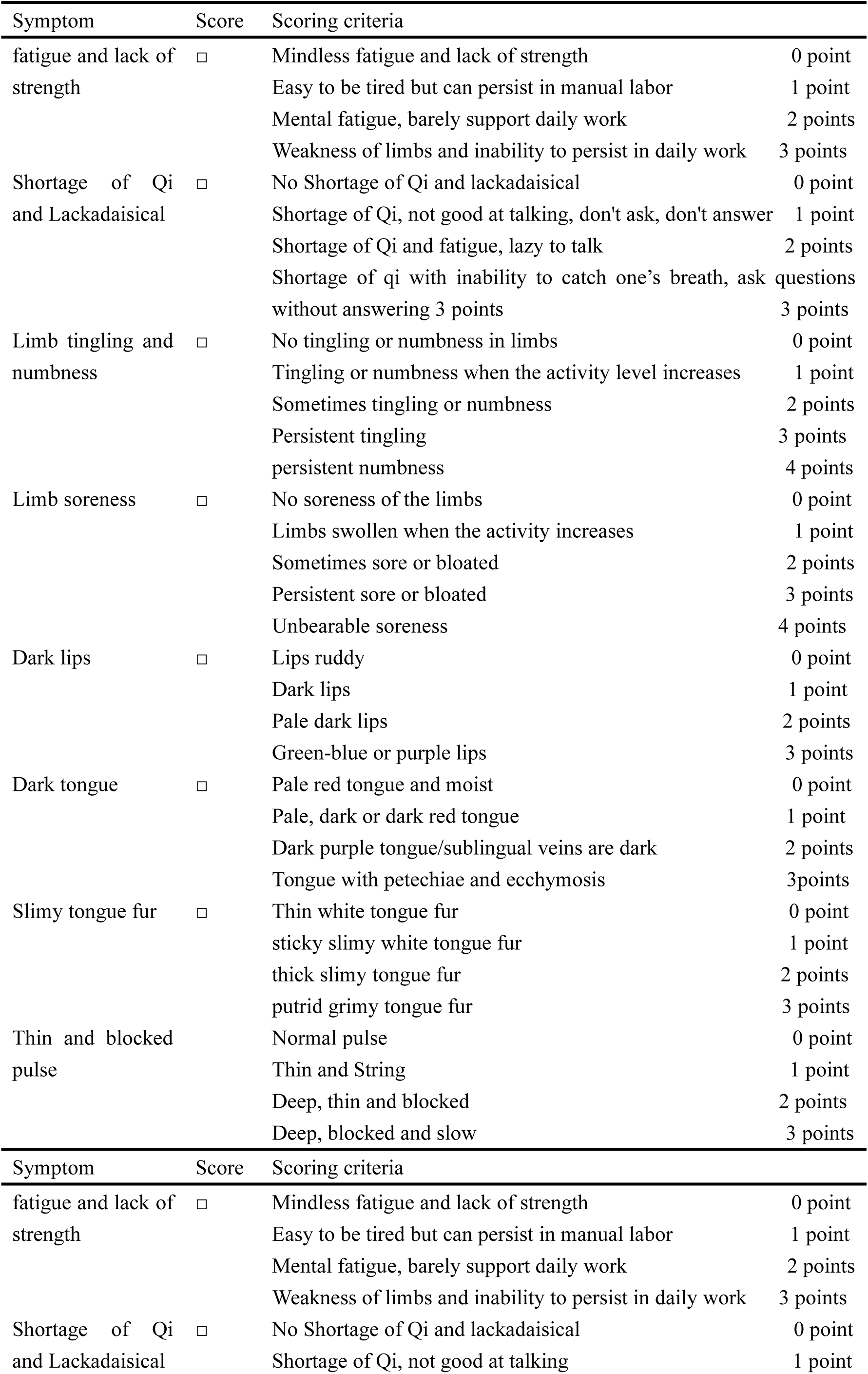
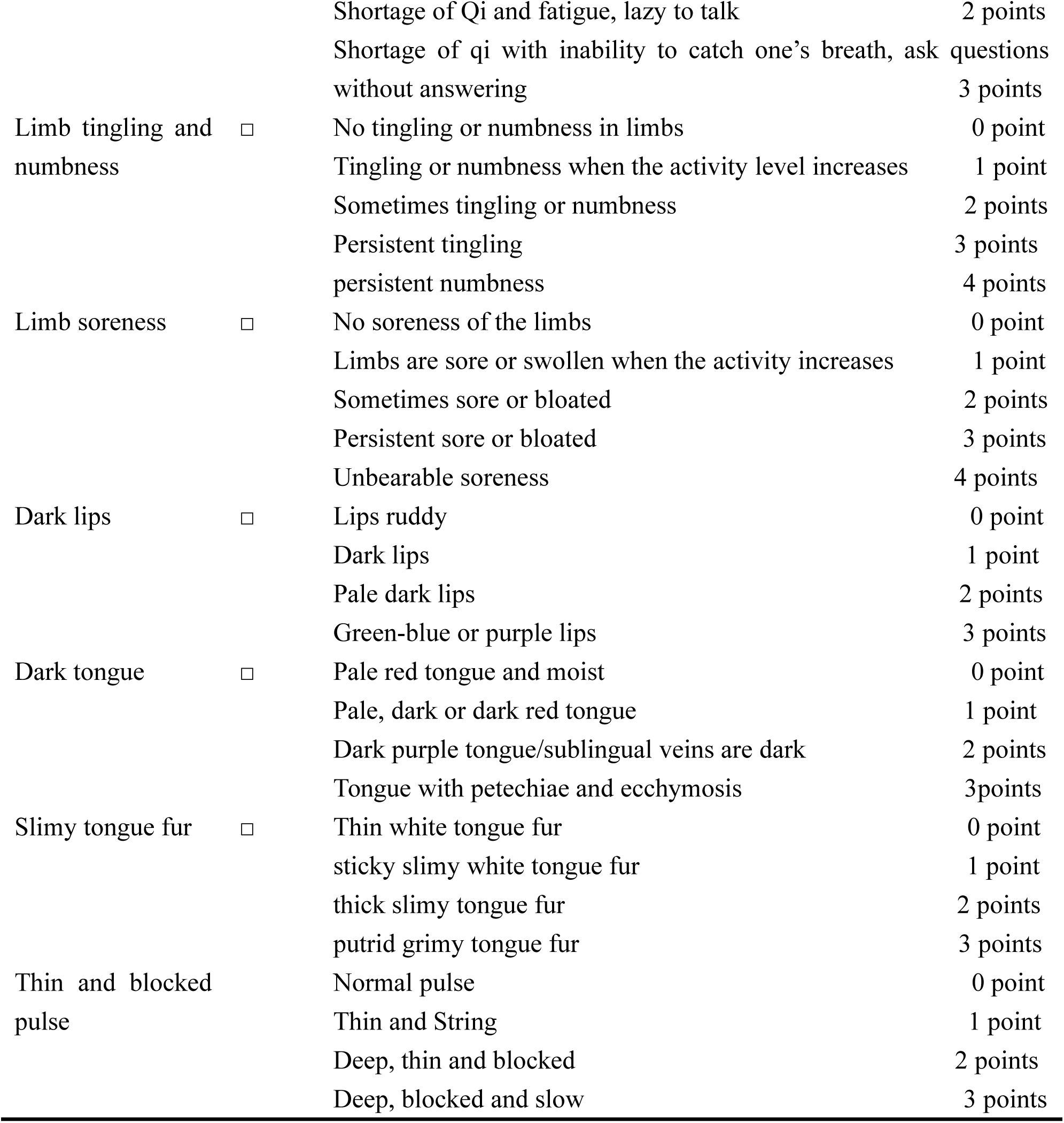
Changes in TCM symptoms scores

